# Metabolic reaction fluxes as amplifiers and buffers of risk alleles for coronary artery disease

**DOI:** 10.1101/2024.08.19.24312222

**Authors:** Carles Foguet, Xilin Jiang, Scott C. Ritchie, Elodie Persyn, Yu Xu, Chief Ben-Eghan, Emanuele Di Angelantonio, John Danesh, Adam S. Butterworth, Samuel A. Lambert, Michael Inouye

## Abstract

Genome-wide association studies have identified thousands of variants associated with disease risk but the mechanism by which such variants contribute to disease remains largely unknown. Indeed, a major challenge is that variants do not act in isolation but rather in the framework of highly complex biological networks, such as the human metabolic network, which can amplify or buffer the effect of specific risk alleles on disease susceptibility. In our previous work, we established that metabolic models can be leveraged to simulate the emerging metabolic effects of genetically driven variation in transcript levels and estimate personalized metabolic reaction fluxes. Here we use genetically predicted reaction fluxes to perform a systematic search for metabolic fluxes acting as buffers or amplifiers of coronary artery disease (CAD) risk alleles. Our analysis identifies 30 risk locus - reaction flux pairs with significant interaction on CAD susceptibility involving 18 individual reaction fluxes and 8 independent risk loci. Notably, many of these reactions are linked to processes with putative roles in the disease such as the metabolism of inflammatory mediators and fatty acids. In summary, this work establishes proof of concept that biochemical reaction fluxes can have non-additive effects with risk alleles and provides novel insights into the interplay between metabolism and genetic variation on disease susceptibility.

## Introduction

Genome-wide association studies (GWAS) have identified tens of thousands of single nucleotide polymorphisms (SNPs) associated with disease risk^1^. However, variant to function (V2F) remains largely unsolved limiting the potential to leverage GWAS results to uncover the underlying disease biology and identify novel therapeutic targets^2,3^. Genetic variants do not act in isolation but within the context of highly complex biological networks that can modulate the effect of specific alleles on disease susceptibility^4,5^. Therefore, unveiling non-additive effects between genetic variants and environment or genetic factors is a powerful approach to understanding the functional relationships of genetic variants and their role in disease^6,7^.

Metabolism is a major biological network comprised of metabolites, enzymes, and transmembrane carriers and underlies many processes in health and disease^8^. One of the most direct manifestations of the metabolic phenotype are metabolic fluxes: the rate at which substrates are converted to products in biochemical reactions or transported across compartments in a metabolic network^9,10^. Dysregulation of metabolic fluxes can play a major role in disease onset and progression^11–14^. For instance, it is well established that alterations in lipid metabolic pathways can disrupt the concentration of lipids in blood and promote the formation of atherosclerotic lesions^15,16^.

Furthermore, metabolic fluxes are particularly attractive as therapeutic targets for perturbation as it has been shown that they can be safely modulated to minimise disease risk and progression. For instance, statins, which are widely prescribed to reduce cardiovascular disease risk, act by reducing the flux of cholesterol synthesis through the inhibition of the enzyme HMG-CoA Reductase^17^. Statins can be combined with inhibitors of bile acid reabsorption, which can further reduce cholesterol levels by increasing the flux of bile acid synthesis^16,18^. Similarly, one of the mechanisms of action of the type 2 diabetes drug metformin, which is also reported to have cardioprotective effects^19^, is the reduction of the oxidative phosphorylation flux^20^.

Notably, intracellular metabolic fluxes cannot be directly measured and instead must be predicted from data integrated into the framework of metabolic networks^9,10^. We have previously demonstrated the feasibility and insights that can be gleaned from using a genotyped population-based biobank to estimate metabolic reaction fluxes and perform a fluxome-wide association analysis for coronary artery disease (CAD)^21^. This analysis was performed using genetically predicted fluxes derived from the integration of genetically imputed transcript abundances^22^ within the constraints defined by the stoichiometric relationships of enzymes and transmembrane carriers in human genome-scale metabolic networks^23^. We found that fluxes through several reactions linked to the metabolism of inflammatory mediators (e.g., histamine and prostaglandins) and polyamines were strongly associated with CAD risk^21^.

Since alterations in metabolic function contribute to disease susceptibility and progression, here we set out to test the hypothesis that metabolic fluxes can also act as buffers or amplifiers of the effects of risk alleles. We performed a systematic search for interactions between genetically personalized organ-specific fluxes and the dosage of risk alleles on CAD susceptibility in UK Biobank (UKB)^24,25^. In doing so, we identified numerous instances where reaction fluxes significantly amplify or buffer the effects of genetic variants on CAD risk.

## Results

### Overview of the methods

We estimated genetically personalized metabolic reaction fluxes for 6,185 organ-specific reactions in 459,902 UK Biobank participants^24,25^ of European genetic ancestries^26^ (**Methods**). From hospital episode statistics and cause of death records for these participants, there were 37,941 CAD cases in total (combined prevalent and incident cases) using the PheWAS Catalog definition of coronary atherosclerosis^27^, and 398,282 non-CAD controls (N=398,282). Using a published genome-wide association meta-analysis performed on over one million participants of European ancestry^28^, we extracted a set of 18,348 SNPs associated with CAD risk (P<5×10^−8^). For downstream analyses, we used the subset of 5,852 SNPs which were found significantly associated with CAD in European UKB participants (P<5×10^−8^; Cox regression model; **Methods**; **Figure 1**).

**Figure 1:**
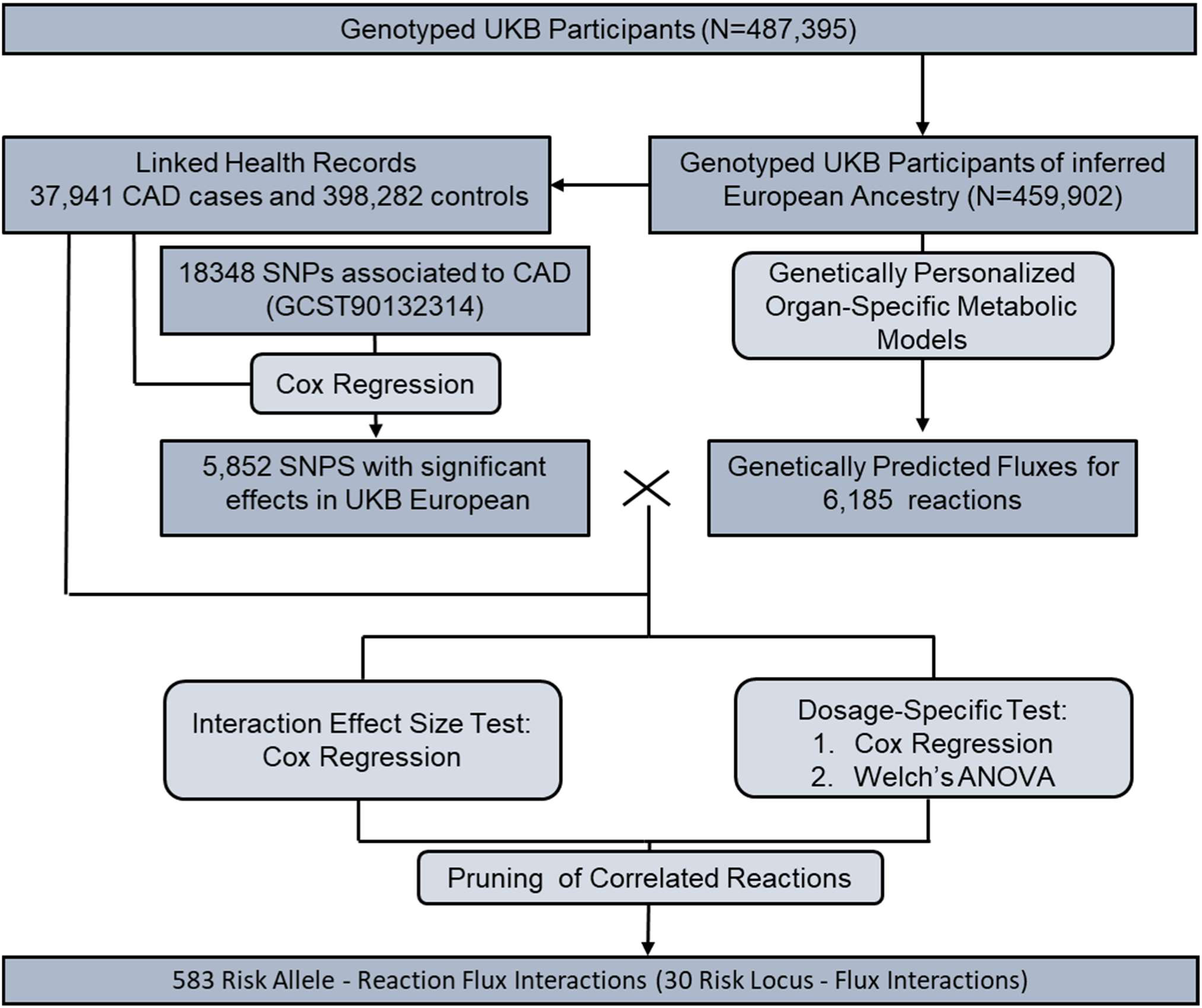
Summary of the methods to detect interactions between reaction fluxes and risk alleles on CAD risk.

We next assessed the extent to which each of the 6,185 reaction fluxes either buffered (i.e. negative interaction) or amplified (i.e. positive interaction) the effects of the 5,852 risk alleles on CAD (**Methods**). To robustly identify such events, we used the intersection of two complementary approaches with different statistical assumptions. In the first approach, we tested for a significant interaction effect size between risk allele dosage and reaction flux value using a Cox proportional-hazards model for CAD events. Before testing for interaction, the effect of the risk allele was regressed from reaction flux to control for potential false positives arising from dependencies between genetically predicted flux values and risk allele dosage^29^. In the second approach, termed dosage-specific test, we tested the differences in reaction flux effect sizes on CAD risk between individuals carrying different risk allele dosages (**Figure S1**; **Methods**). The P-values from the interaction model and the dosage-specific test were then adjusted for multiple testing using the Benjamini-Hochberg method (i.e. FDR). A buffering or amplification effect of a reaction flux on a risk allele had to be significant under both approaches (FDR-adjusted P-value <0.05; **Figure 1**) to be deemed valid.

### Buffering and amplification of the effect of risk variants by reaction fluxes

In total, we found 583 pairs of SNP-reaction fluxes which were significant (FDR-adjusted P-value <0.05) in both the interaction effect size test and the dosage-specific test (separately, 669 and 595 SNP-reaction flux pairs were significant, respectively) (**Supplementary Data S1**). We observed strong correlations between interaction effect estimates (r= 0.998) and the p-values (r=0.713) of both approaches (**Figure S2**). Indeed, from the 86 pairs significant for the interaction effect size test but not the dosage-specific test, 82 were significant with an FDR-adjusted P-value<0.25 in the latter. Similarly, 7 out of the 12 pairs significant with the dosage-specific test but not the interaction effect size test were also borderline significant, with the remaining pairs being instances where the interaction might significantly deviate from linearity.

Of the 583 SNP-reaction flux pairs, 353 displayed buffering (i.e. negative interaction effect size) and 230 displayed amplification (i.e. positive interaction effect size) of the effects of the risk allele by reaction fluxes(**Figure 2**). The significant pairs comprised 279 unique SNPs mapped to 8 independent risk loci (R^2^<0.6; **Methods**) leading to 30 risk locus – reaction flux pairs with significant interaction on disease susceptibility. These interactions encompassed 18 unique reaction fluxes. Notably, only five of these fluxes had a significant effect on CAD risk in univariate association analysis, indicating that the majority of the metabolic associations here unveiled have their association with CAD masked unless analysed together with common risk alleles(**Figure S3**).

**Figure 2:**
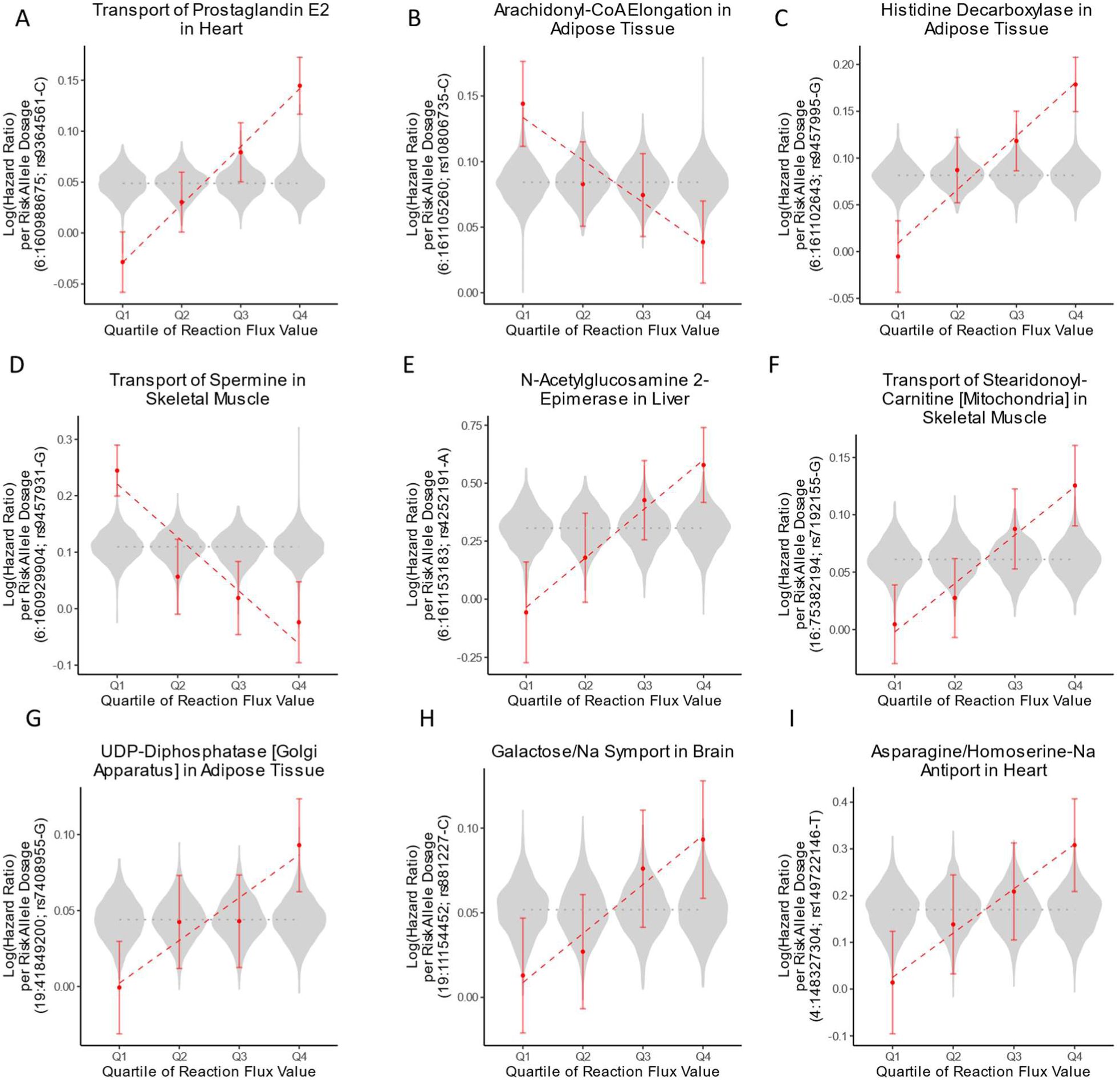
Examples of buffering and amplification of variant effect sizes by reaction fluxes on CAD risk. For each reaction, UKB participants of European genetic ancestries were quartile-binned according to the personalized reaction flux value, and variant effect sizes (Log(Hazard Ratio)) were estimated within each subset using Cox regression. Error bars denote the 95% confidence intervals for variant effect sizes. The dashed lines indicate the linear regression of variant effect sizes per flux quartile. Violin plots indicate the distribution of variant effect size on CAD risk for all other reaction fluxes. The dotted grey line indicates variant effect size in all UKB participants of European genetic ancestries. Genome coordinates correspond to the GRCh37 genome assembly.

### The genomic region encoding Lp(a) and plasminogen is a major site for amplification and buffering

The majority (530 out of 583) of the SNP-reaction flux pairs with significant interaction on CAD risk were mapped to four risk loci in the chromosome 6 genomic region encoding the *LPA* and *PLG* genes (**Figure 2A-E; Figure 3A; Figure S1A-E; Figure S4; Supplementary Data S1**). *LPA* codes for apolipoprotein(a) which is the primary constituent of Lp(a) and has been established to have a causal role in the formation of atherosclerotic lesions by promoting lipid accumulation, inflammation, and calcification in the artery wall^30–32^. *PLG* encodes plasminogen which can contribute to atherosclerosis by modulating, cell migration, extracellular matrix structure, vascular smooth muscle cell (VSMC) function, and inflammation^33–35^.

**Figure 3:**
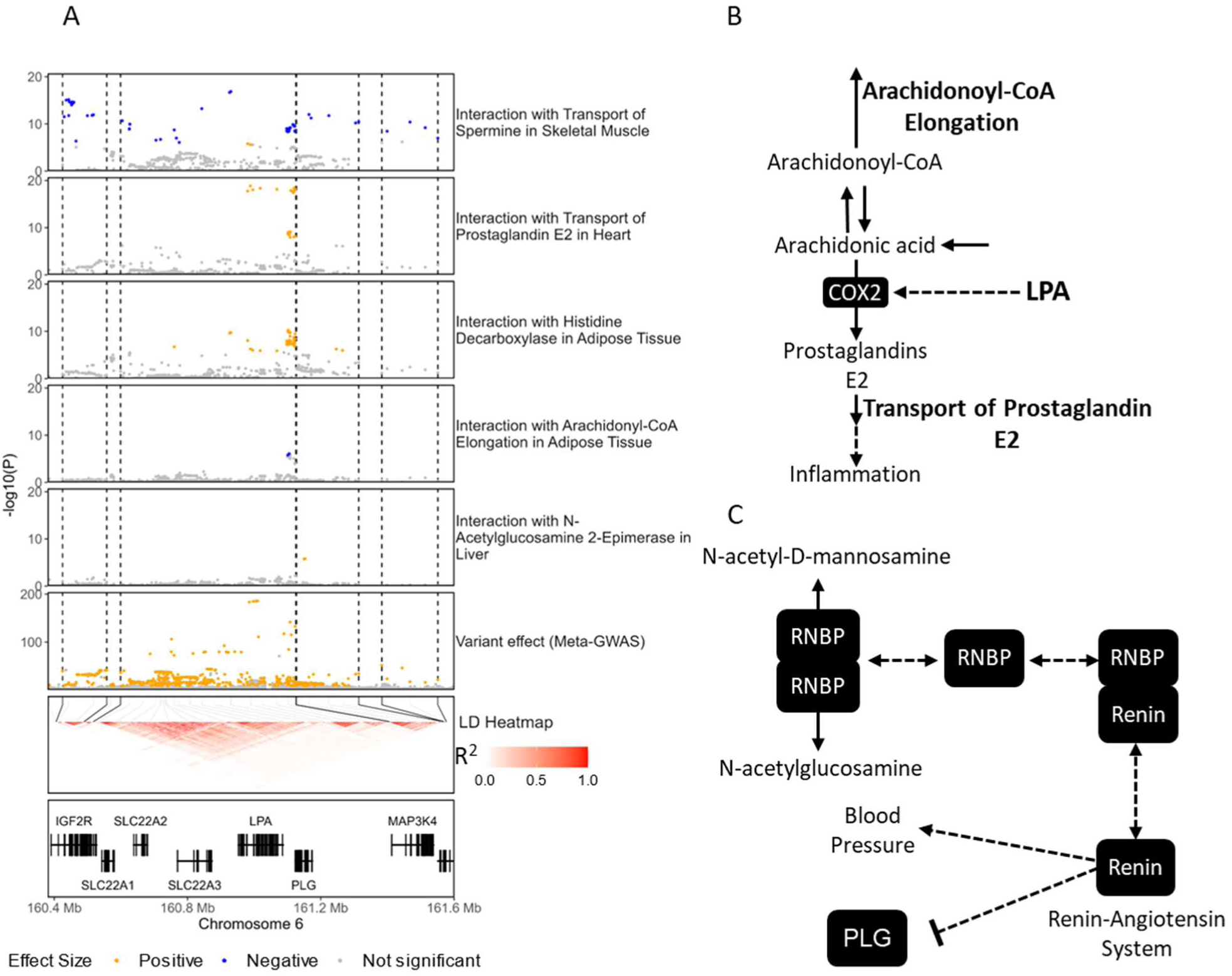
Examples of amplification and buffering at the *LPA/PLG* risk loci. A) Regional association plots showing the −log10(P-value) for interaction and variant effect sizes on CAD risk. P-values for interaction and variant effect sizes were derived from the interaction effect size test and meta-GWAS summary statistics, respectively. Only a representative subset of reactions involved in significant interactions are shown, the complete set of interactions for this region is shown in **Figure S4**. The LD heatmap indicates the pairwise LD for SNPs with genome-wide significant effect size on CAD in UKB participants of European genetic ancestries. Dashed black lines indicate the limits of LD blocks (R^2^>0.6) used to define independent risk loci. To facilitate visualization, only LD blocks with variants involved in significant interactions are highlighted. Only protein-coding genes are shown in the gene plot. Genome coordinates correspond to the GRCh37 genome assembly. B) Potential mechanism of interaction between reactions of prostaglandin metabolism and *LPA*. LPA induces the expression of the enzyme COX2 which catalyses the formation of prostaglandin E2 from arachidonic acid. Conversely, the elongation of arachidonoyl-CoA diverts arachidonic acid away from prostaglandin synthesis. C) Potential mechanism of interaction between N-Acetylglucosamine 2-Epimerase and plasminogen. This reaction is catalysed by a homodimer of RNBP, which can also form a heterodimer with renin inhibiting the renin-angiotensin system. The renin-angiotensin system can suppress plasminogen activation. Solid arrows denote metabolic reactions or transport processes and dashed lines other functional relationships.

The flux of prostaglandin E2 transport in both heart and brain tissues strongly amplifies the effect size of a set of risk variants mapped to the risk locus which includes the *LPA* gene(**Figure 2A; Figure 3A; Figure S1A; Figure S4**). Prostaglandin E2 is an inflammatory mediator that plays a role in the pathogenesis of atherosclerosis^36^. Interestingly, the flux of elongation of arachidonoyl-CoA in adipose tissue has the opposite effect and buffers the effect size of two SNPs in the same risk locus (**Figure 2B; Figure S1B**). Arachidonoyl-CoA is the CoA-conjugated form of arachidonic acid which is a precursor for prostaglandin synthesis^23^ (**Figure 3B**). Similarly, variants in the same locus also had their effect size amplified by the flux of histamine synthesis in adipose tissue (**Figure 2C; Figure S1C**) and a flux of histamine transport into the liver(**Figure S4**). Histamine is also an inflammatory mediator that has been linked to atherosclerosis by modulating inflammation, blood lipids, and lipoprotein fractions^37,38^.

We also found that fluxes involving polyamine transport in adipose tissue, heart, and skeletal muscle can have buffering or amplification effects in a variant- and tissue-specific manner across the four adjacent risk loci (**Figure 3A; Figure 2D; Figure S1D; Figure S4**). Polyamines are a family of pleiotropic compounds that regulate cell proliferation, cell differentiation, and protein synthesis and have a generally protective effect against inflammation and oxidative stress^39^. Polyamine-rich diets have been established to protect against cardiovascular disease by countering the age-related myocyte and vascular endothelial dysfunctions^39^ while dysregulation of endogenous polyamine metabolism can also lead to pathologies such as cardiac hypertrophy^40,41^. In adipose tissue, polyamine synthesis is reported to protect against obesity by promoting vascularization and lipolysis^42^ while in skeletal muscle it promotes muscle mass growth and regeneration^43^.

A commonality between prostaglandin and polyamine transport is that they are mediated by transmembrane carriers coded by genes in the *LPA*/*PLG* genomic region. Namely, prostaglandin transport can be mediated by either *SLC22A1*, *SLC22A2*, or *SLC22A3* and polyamine transport is mediated by *SLC22A1*^23,44^. In this regard, expression quantitative trait loci (eQTL) variants for these transporters had been used as input for computing genetically personalized flux values^21,22^ (**Methods**) and are also mapped within this genomic region. In addition to regressing out the effect of the individual risk allele when testing for interaction, we quantified the linkage disequilibrium (LD) between eQTL variants used in flux computation and CAD risk variants, and the contribution of the former to flux values. We found that, while some eQTL variants had a strong contribution to reaction fluxes, they were not in strong LD with CAD risk variants with significant interaction with reaction fluxes(**Figure S5**). Furthermore, the flux through the elongation of arachidonoyl-CoA was not associated with any eQTL variants at the *LPA/PLG* loci, suggesting that prostaglandin metabolism may modulate the effect of risk alleles independently of the activity of the SLC22A1*-3* transporters.

Finally, in the risk locus encoding the *PLG* gene, two intron variants displayed significant interaction on disease risk with the flux through N-Acetylglucosamine 2-Epimerase in the liver(**Figure 3A; Figure 2E; Figure S1E**). This reaction is catalysed by a homodimer of the renin-binding protein (RNBP)^23,45^. The formation and stabilization of its catalytically active form prevents the formation of a heterocomplex with renin, which sequesters the latter and inhibits the renin-angiotensin system^45^ (**Figure 3C**). The renin-angiotensin system increases blood pressure and has been linked to atherosclerosis^46^.

### Transport of stearidonoyl-carnitine amplifies the effect size of variants mapped to the *BCAR1/CFDP1* risk locus

In the *BCAR1*/*CFDP1* risk locus, there was evidence of risk allele amplification by the flux of mitochondrial transport of stearidonoyl-carnitine in skeletal muscle, with 38 risk variants showing significant interaction with this flux(**Figure 2F; Figure 4A; Figure S1F; Supplementary Data S1)**. Polymorphisms within this region have been associated with carotid intima-media thickness (i.e. a marker of subclinical atherosclerosis) and CAD risk with *BCAR1* identified as the likely causal gene^47,48^. *BCAR1* regulates cell migration, proliferation and apoptosis and is essential for cardiovascular development in embryogenesis^49^. In particular, *BCAR1* is a major regulator of VSMC function and it has been theorized that *BCAR1*’s contribution to the formation of atherosclerotic lesions arises from its role in VSMC migration^50^.

**Figure 4:**
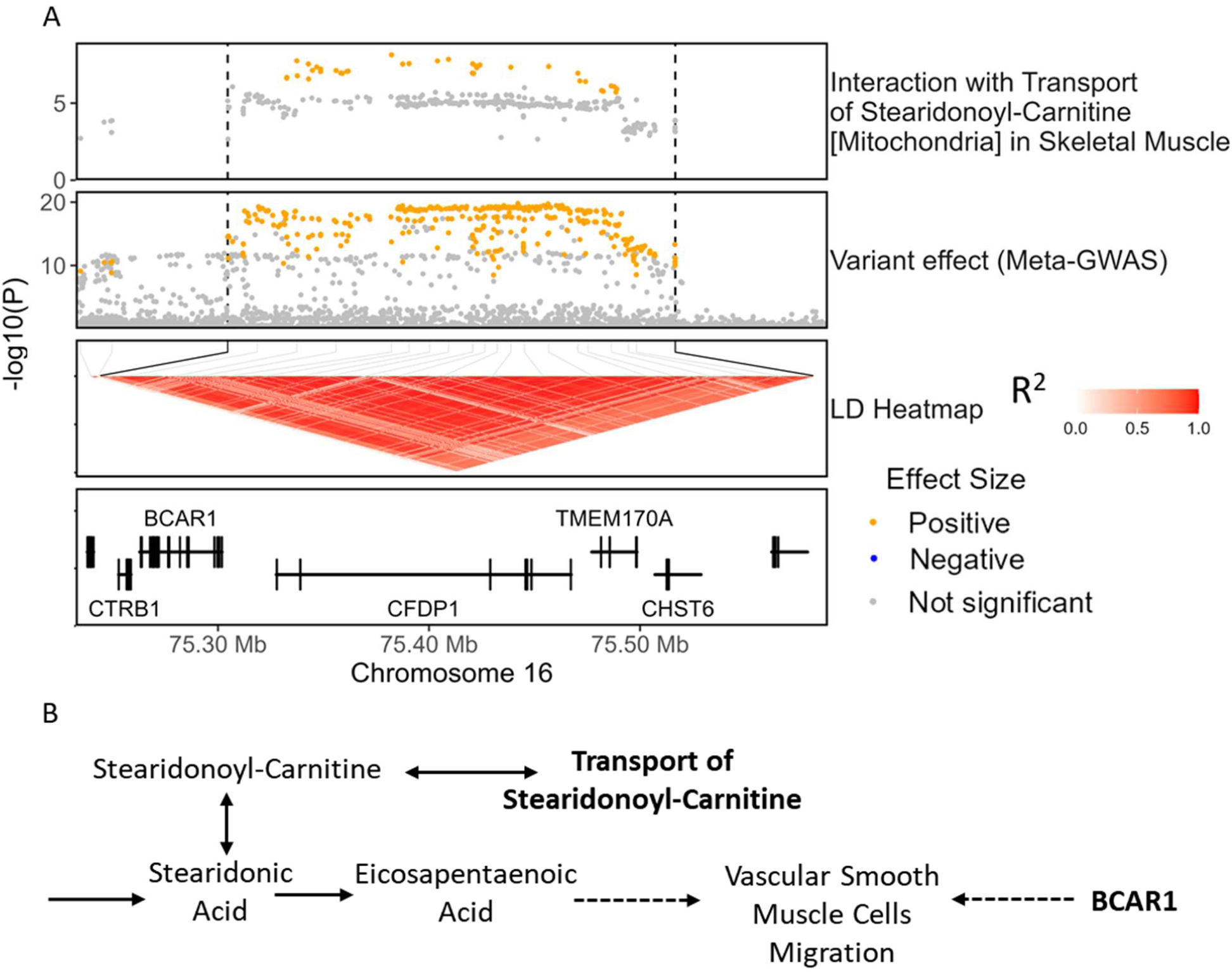
Transport of stearidonoyl-carnitine amplifies the effect size of risk variants of the *BCAR1*/*CFDP1* locus. A) Regional association plots showing the −log10(P-value) for interaction and variant effect sizes on CAD risk. P-values for interaction and variant effect sizes were derived from the interaction effect size test and meta-GWAS summary statistics, respectively. B) Potential mechanism of interaction between the transport of stearidonyl-carnitine and risk variants linked to *BCAR1*. Solid arrows denote metabolic reactions or transport processes and dashed lines other functional relationships, namely activation of the migration of vascular smooth muscle cells.

Stearidonoyl-carnitine is the carnitine-conjugated form of stearidonic acid (SDA), an omega-3 polyunsaturated fatty acid that can be efficiently metabolized to eicosapentaenoic acid (EPA)^51^. EPA has well-established anti-inflammatory and cardioprotective effects^52,53^ and has been shown to inhibit the progression of atherosclerotic lesions^54^. Likewise, SDA has also been found to have anti-inflammatory effects in cell and animal models^51,55^. However, SDA levels in the blood have also been associated with an increased risk of cardiovascular pathologies such as hypertension^56^ or thrombosis^57^ suggesting a context-specific role in disease.

### Galactose transport into the brain amplifies the effect size of variants mapped to the *SMARCA4* risk locus

A set of 19 variants mapped to a risk locus encoding the *SMARCA4* gene, were identified as having their effect size amplified by the flux of the sodium-coupled galactose transport in the brain(**Figure 2H; Figure S1H; Figure S6; Supplementary Data S1)**. *SMARCA4* codes for a chromatin-remodelling factor that has a major role in transcriptional regulation, DNA repair, and cell proliferation in a wide range of processes and tissues^58^. The *SMARCA4* gene is adjacent to *LDLR*, a well-established risk locus for CAD, but it has LDLR-independent roles in CVD risk^59^. For instance, *SMARCA4* has been reported to mediate vascular calcification^60^, inflammation^59^ and myocardial proliferation^61^. Indeed, the variants showing significant interaction with galactose transport are in a different LD block than those linked to *LDLR* (**Figure S6**).

### UDP-Diphosphatase amplifies the effect size of variants mapped to the TGF-β-risk locus

In the TGF-β risk locus, there was evidence of risk allele amplification by the flux through UDP-diphosphatase (Golgi apparatus) in adipose tissue, with two intron variants of *TGFB1* showing significant interaction on disease risk with this flux (**Figure 2G; Figure 5A; Figure S1G; Supplementary Data S1)**. *TGFB1* codes for a ligand of the Transforming Growth Factor β (TGF-β) family which regulates a wide range of biological processes such as morphogenesis, tissue homeostasis, and inflammation in a context-specific manner^62^. TGF-β plays a major role in the cardiovascular system by regulating the proliferation, differentiation and function of endothelial, smooth muscle, and immune cells, and its dysregulation can lead to a wide range of cardiovascular diseases such as atherosclerosis^63^. TGF-β signalling is also linked to adiposity through the regulation of adipocyte differentiation and oxidative metabolism^64^, and *TGFB1* is overexpressed in obesity^65,66^.

**Figure 5:**
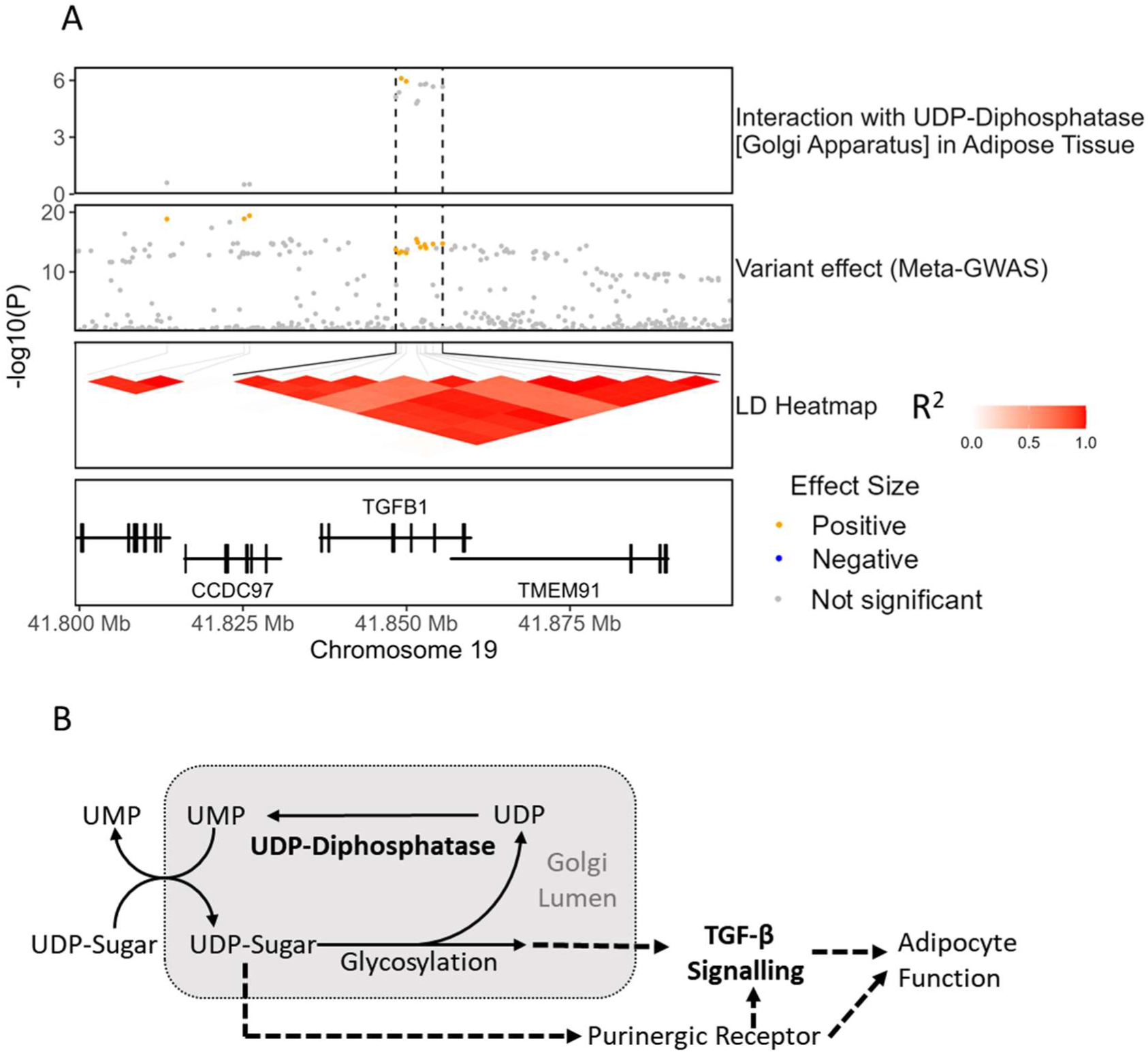
UDP-diphosphatase amplifies the effect size of CAD risk variants within the *TGF-β* risk locus. A) Regional association plots showing the −log10(P-value) for interaction and variant effect sizes on CAD risk. P-values for interaction and variant effect sizes were derived from the interaction effect size test and meta-GWAS summary statistics, respectively. B) Potential mechanism of interaction between UDP-diphosphatase and risk variants linked to *TGB1*. Solid arrows denote metabolic reactions or transport processes and dashed lines other functional relationships.

UDP-diphosphatase plays a major role in the Golgi apparatus, which is the primary location of protein and lipid glycosylation. Nucleotide sugars (such as UDP-sugars) are transported from the cytosol to the Golgi’s lumen where they act as donors for a glycosylation process releasing the nucleotide diphosphate (e.g. UDP)^67^. Notably, the transport of UDP-sugars to Golgi involves an antiport with luminal UMP^68,69^. As such, UDP-Diphosphatase contributes to glycosylation by catalysing the hydrolysis of UDP to UMP hence enabling the UMP-dependent transport of nucleotide sugars^67,68^. Additionally, the UMP-dependent transport of nucleotides-sugars to the Golgi’s lumen is also reported to contribute to the vesicle-based release of UDP-sugars to the extracellular space^69^ (**Figure 5B**).

### Amino acid transport amplifies the effect of a risk variant upstream of the *EDNRA* gene

A significant positive interaction in CAD risk was detected between a risk variant, mapped upstream of the *EDNRA* gene, and the flux of an amino acid transport process in the heart (i.e. the sodium-coupled exchange of homoserine and asparagine) (**Figure 2I; Figure S1I; Figure S7; Supplementary Data S1)**. *EDNRA*, which is expressed in VSMC and cardiomyocytes, codes for the receptor for endothelin-1. Endothelin-1, acting through the activation of EDNRA, is a potent vasoconstrictor with a well-established role in cardiovascular diseases^70^. *EDNRA* expression in vascular tissue and endothelin-1 levels in blood are altered in atherosclerosis^71,72^ and EDNRA antagonists can attenuate the progression of coronary atherosclerosis lesions^73^. The concentration in blood of both asparagine and homoserine have been associated with cardiovascular disease risk^74–77^, however the mechanism of their effect remains largely unknown.

### Buffering and amplification of the risk of myocardial infarction

Myocardial infarction (MI) is caused by a sudden blockage of blood flow to the myocardium, primarily due to coronary atherosclerosis with or without a blood clot^78^. Given the close relationship between MI and CAD and that the metabolic reactions we assess here may not *a priori* be involved in hard outcomes like MI, we investigated the extent to which the identified instances of buffering and amplification of risk allele penetrance on CAD risk could also be relevant for MI risk.

On the UKB participants of European genetic ancestries, we identified 36,007 MI cases and 423,629 controls using an established definition of MI^79^. As expected, there was a substantial overlap of cases and controls between MI and coronary atherosclerosis (**Table S1**). Risk variant and reaction flux effect sizes, estimated using Cox regression, were highly correlated between coronary atherosclerosis and MI (**Figure S8A-D**). However, reaction effect sizes for MI risk were on average lower than the equivalent effect sizes derived for coronary atherosclerosis.

We evaluated buffering and amplification effects between risk variants and risk allele dosage on MI risk. We tested the same SNP-reaction flux pairs that had been evaluated with coronary atherosclerosis to facilitate the comparison of the results. For MI, there were 426 pairs significant with both interaction effect size and dosage-specific tests (FDR-adjusted P-value<0.05) **(Supplementary Data S1)**. Taking into account the LD structure of risk SNPS, these represented 26 risk locus – reaction flux pairs with evidence of significant amplification or buffering of risk alleles by reaction fluxes. Out of the 583 significant SNP-flux pairs for coronary atherosclerosis, 360 were also significant in MI (20 risk locus – reaction flux pairs) with 91 additional SNP-flux pairs (4 additional risk locus – reaction flux pairs) being borderline significant (FDR-adjusted P-value<0.25 for both the interaction effect size test and the dosage-specific tests). Overall, interaction effect sizes were also strongly correlated between both MI and coronary atherosclerosis (**Figure S8E-H**).

Notably, there were no significant interactions on MI risk between the transport of stearidonoyl-carnitine and the risk locus of *BCAR1*/*CFDP1* nor UDP-diphosphatase and the *TGFB1* risk locus. Effect sizes for variants mapped to these loci estimated with Cox regression were also slightly lower in MI compared to coronary atherosclerosis (**Figure S9**). Conversely, two loci had significant buffering of disease risk alleles by reaction fluxes in MI and not in CAD: effect sizes of 15 risk alleles mapped to the *PDE5A*/*MAD2L1* locus, and 13 risk alleles mapped to the *MAP1S/FCHO1* locus were buffered by the flux of orotate phosphoribosyltransferase in heart and uracil transport in skeletal muscle, respectively (**Figure S10**). Both reactions are functionally part of pyrimidine metabolism, which has been implicated in MI^80–82^. Notably, PDE5A is also linked to nucleotide metabolism and its inhibition is well-established to have cardioprotective effects in MI^83,84^.

## Discussion

Here, we used genetically personalized organ-specific metabolic fluxes to study the interaction between risk variants and biochemical reactions on disease risk using CAD as a case study. Our analysis identified 18 metabolic reaction fluxes that can amplify or buffer risk allele effect sizes at 8 well-established CAD risk loci unveiling a total of 30 risk locus - reaction flux pairs with significant interaction on disease susceptibility. Most of such reactions involved metabolic processes with known roles in atherosclerosis such as inflammation or fatty acid metabolism. Furthermore, the majority of the interactions detected for CAD were also relevant for MI.

We identify that the genomic region of *LPA*/*PLG*, a well-known locus for CAD risk^28,85^, is a major site of interaction between risk variants and reaction fluxes. For instance, we find that a set of variants in the *LPA* and *PLG* risk loci have their effect size amplified by the flux of reactions involved in the synthesis or transport of histamine and prostaglandin E2, two inflammatory mediators that can contribute to the formation of atherosclerotic lesions^36,38^. In our previous work^21^, we identified that some of those reaction fluxes were associated with CAD risk, here we show that this effect can be further amplified or buffered by the dosage of specific risk alleles within the *LPA*/*PLG* region (**Figure S1A,C**). Inflammation is one of the mediators of pathogenicity of *LPA* and *PLG*^31–33^ and the former has been reported to induce the expression of cyclooxygenase-2, which catalyses the first step of prostaglandin E2 synthesis^31^. Hence, the identified interactions may reflect a mechanism where *LPA or PLG* variants that increase inflammation have their effect on CAD risk amplified by a high capacity to transport prostaglandin E2 or histamine across cellular membranes. Conversely, the flux of elongation of arachidonoyl-CoA in adipose tissue would attenuate this effect by diverting arachidonic acid away from prostaglandin synthesis (**Figure 3B**).

Notably, most interactions were also instances where the effect of a biochemical reaction flux on CAD risk only becomes apparent when analysed in conjunction with risk variants. For instance, neither the flux through N-Acetylglucosamine 2-Epimerase in the liver, the transport of stearidonoyl-carnitine in skeletal muscle nor UDP-diphosphatase in adipose tissue have a significant effect on disease risk when analysed in univariate analysis. However, the interaction analysis reveals that such fluxes can significantly amplify the effect size of risk variants mapped to the *PLG, BCAR1* and *TGFB1* risk loci, respectively, providing insights into their roles in cardiovascular disease. For instance, the reaction N-Acetylglucosamine 2-Epimerase is mediated by an enzyme that moonlights as an inhibitor of renin^45^ and the formation of its catalytically active form prevents it from inhibiting the renin-angiotensin system^46^, a known regulator of plasminogen^86,87^, suggesting a potential mechanism of interaction with variants of the plasminogen risk locus(**Figure 3C**). Concerning the transport of stearidonoyl-carnitine, the transport of acylcarnitines to mitochondria is the limiting step for mitochondrial β-oxidation^88^, and hence this reaction might be affecting CAD risk by modulating the bioavailability of SDA and other polyunsaturated fatty acids. In this regard, one of the cardioprotective actions of omega-3 polyunsaturated fatty acids is the inhibition of VSMC proliferation and migration^52,89^, thus providing a link to *BCAR1* which is also reported to modulate VSMC function^50^ (**Figure 4B**). Similarly, UDP-diphosphatase is reported to play a major role in the transport of nucleotide-sugars to the Golgi apparatus^67,68^ and hence can potentially modulate both protein glycosylation and the vesicle-based release of UDP-sugars^69^. The formation of the functional form of TGF-β, and many of the other proteins involved in TGF-β signalling, requires glycosylation in the Golgi apparatus^90,91^ suggesting a potential avenue of interaction. Additionally, the release of UDP-sugars, acting through the activation of purinergic receptors, can modulate adipocyte differentiation, lipolysis, and inflammation within the adipose tissue^92^. Indeed, there is ample evidence of crosstalk between TGF-β signalling and purinergic receptors^93–95^ suggesting a second potential mechanism of interaction (**Figure 5B**).

In conclusion, this work establishes proof of concept that biochemical reaction fluxes can modulate the effect of disease risk alleles and highlights the importance of considering the burden of risk variants to understand the contribution of metabolism to cardiovascular disease susceptibility. Given that disease-associated metabolic processes represent potential targets against disease, such findings have important implications for personalized medicine as they highlight that the therapeutic efficacy of targeting specific metabolic pathways may depend on each individual’s genetic background.

## Methods

### UK Biobank

UKB is a cohort of approximately 500,000 participants from the general UK population (https://www.ukbiobank.ac.uk/). Participants were between age 40 and 69 at recruitment (median 58 years of age; 54% women) and accepted an invitation to attend one of the assessment centres that were established across the United Kingdom between 2006 and 2010^24^. We used the version 3 release of the UK Biobank genotype data^25^ (https://biobank.ndph.ox.ac.uk/showcase/label.cgi?id=263), which had been imputed to the UK10K/1000 genomes and haplotype reference consortium (HRC)^96^ panels.

### Ancestry inference

Genotyped UKB participants were assigned a genetic ancestry using KING^26^. Briefly, genotyped UKB samples were projected to the 10 main genetic principal components computed from samples from the 1000 genome project with known superpopulation groups (American, East Asian, European, and South Asian). Using this projection, KING uses a support-vector-machine-based method to infer the most likely ancestral group of each sample^26^. UKB participants were assigned to European Ancestry if the probability of belonging to that group was estimated to be 95% or higher.

### Genetically personalized organ-specific fluxes

Genetically personalized organ-specific fluxes were computed as previously described^21^. Briefly, organ-specific metabolic models for adipose tissue, brain, heart, liver and skeletal muscle were extracted from Harvey/Harvetta whole body models^97^ and ported to HUMAN1^98^, the latest reconstruction of human metabolism. The GIM3E^99^ algorithm was then used to compute an average reaction flux distribution for each organ consistent with average organ-specific transcript abundances obtained from GTEx^100^. In parallel, genotype data was used to impute personalized organ-specific transcript abundances for UKB participants using the elastic net models from PredictDB^22^. Finally, the quadratic metabolic transformation algorithm (qMTA) was used to integrate the individual-level organ-specific transcript abundances and the average reaction flux distribution and compute genetically personalised organ-specific reaction flux values for each analysed organ. Flux values for each reaction were log2-transformed and standardized to zero-mean and unit variance.

### Definition of coronary artery disease

Cases of CAD were identified using the definition of coronary atherosclerosis (Phecode 411.4) of the PheWAS Catalog (version 1.2)^27^. Namely, cases were defined by the presence of any of the constitutive ICD9 (411.81, 414.0, 414.01, 414.02, 414.03, 414.04, 414.05, 414.2, 414.3, 414.4, 996.03 or V45.81, V45.82) and ICD10 (I24.0, I25.1, Z95.1 or Z95.5) codes in hospital episode statistics or death records. Additionally, non-cases with any of the constitutive ICD codes of ischemic heart disease (Phecode 410-414.99) were excluded from the controls^27^. We identified 37,941 CAD cases and 398,282 controls in genotyped UKB participants of European ancestry. The earliest coded or reported date for disease was converted to the age of phenotype onset. Controls were censored according to the maximum follow-up of the health linkage data (October 31, 2022) or the date of death.

### Definition of myocardial infarction

MI was defined as evidence of a fatal or nonfatal myocardial infarction or major coronary surgery in hospital episode statistics, death records, or self-reported during the verbal interview at UKB enrolment^79^. More in detail, myocardial infarction was defined as the presence of ICD-9 codes 410-412, ICD-10 codes I21–I24 or I25.2 in hospital episode statistics or cause of death records or reporting a heart attack during the verbal interview at UKB enrolment (Self-report field 6150 and Self-report field 20002). Major coronary surgery was defined by ICD-9 code V45.81, ICD-10 code Z95.1, OPCS-3 codes 309.4 or 884, OPSC-4 codes K40–K46 or reporting a coronary angioplasty, coronary artery bypass grafts or triple heart bypass at the verbal interview (Self-report field 20004)^79^. We identified 36,007 cases and 423,629 controls in genotyped UKB participants of European ancestry. The earliest coded or reported date for disease was converted to the age of phenotype onset. Controls were censored according to the maximum follow-up of the health linkage data (October 31, 2022) or the date of death.

### Risk variant selection

A set of variants associated with CAD risk was obtained from a published genome-wide association meta-analysis performed on over one million participants of European ancestry^28^. Summary statistics from this study were downloaded from the GWAS catalog (GCST90132314)^1^ and they identified 18,348 biallelic SNPs with genome-wide significance (P<5×10^−8^).

We anticipated that only a subset of these variants might have significant effects in our cohort. As such, we tested the effect of these variants on CAD risk in the European subset of UKB. For this, we used a similar model and assumptions that we would subsequently use for the amplification and buffering analysis (i.e., Cox proportional-hazards model using age as a time scale stratifying by sex and genotyping array and using the first 10 genetic principal components as covariates). We selected the 5,852 variants that had genome-wide significance in our cohort as candidate variants to test for amplification and buffering of disease risk by reaction fluxes.

### Identifying interactions between reaction fluxes and risk variants on disease risk

To robustly identify instances of buffering/amplification between reaction fluxes and risk variants we used two complementary methods to detect interaction: an interaction effect size test and a dosage-specific test.

In the interaction effect size test, for each pair of reaction flux values and risk allele dosages, we test for a significant interaction term using a Cox proportional-hazards model with age as a time scale for CAD risk:

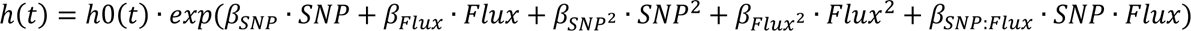

where,

ℎ(*t*) is the hazard function defining the risk of CAD at age *t*

ℎ0 is the baseline hazard

*SNP* is the dosage of the risk allele

*Flux* is the adjusted reaction flux value. Before testing for interaction, flux values are adjusted with linear regression to remove any potential effects of *SNP* over the reaction flux value.

*β_SNP_*, *β_Flux_*, *β_SNP2_*, *β_Flux2_* are the first and second-order effect sizes for SNP dosage and Flux value *β_SNP:Flux_SNP* · *Flux* is the interaction effect size between SNP dosage and Flux value The first ten genetic principal components were also used as covariates but have been omitted from this equation for clarity. Additionally, this analysis was stratified by sex and genotyping array. The model was fitted using the CoxPHFitter function from the lifelines python package^101^. The significant interaction effect on CAD risk was evaluated with a two-tailed Wald test for the interaction effect size.

In the dosage-specific effect size test, for each pair of reaction flux values and risk alleles, we split the analysed UKB participants based on risk allele dosage (0, 1 or 2) and estimated the effect of reaction flux value on CAD risk within each allele dosage using a Cox proportional hazards model with age as time scale.

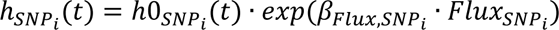

where,

ℎ*_SNP_i__*(*t*) is the hazard function defining the risk of developing CAD at age *t* in individuals with dosage *i* of the risk allele

ℎ0*_SNP_i__* is the baseline hazard in individuals with dosage *i* of the risk allele

*β_Flux,SNP_i__* is flux effect size in individuals with dosage *i* of the risk allele.

*Flux* is the reaction flux value. Adjusting flux values to regress out any potential effects of *SNP* dosage has no effect in this analysis as the dosage of the risk allele is constant for each test.

As with the interaction model, the first ten genetic principal components are also used as covariates and the analysis is stratified by sex and genotyping array.

The CoxPHFitter function from the lifelines python package^101^ was used to estimate the effect size of reaction flux values and its standard error (SE) for each dosage of the risk allele. A Welch’s ANOVA test (i.e. a variant of ANOVA that does not assume homogeneity of variance)^102^ was used to evaluate if there were significant differences between reaction effect sizes across risk allele dosages. To facilitate comparing these results with the interaction effect size, effect size variation per dosage of risk allele were computed with a linear regression of flux effect size per dosage weighted by the standard error of effect sizes estimates at each dosage (1/SE^2^).

### Reaction selection and pruning

In the HUMAN1-derived organ-specific models^21,23^, some reactions lack gene annotation. This can occur because they are spontaneous processes, but more often than not this arises because the gene(s) mediating the reaction are unknown or because they are artificial reactions needed to balance the models (e.g., exchange reactions). As such, we decided to focus the amplification/buffering analysis only on the fluxes of reactions with annotated genes (adipose tissue: 721, brain: 1116, heart: 1314, liver: 1956, skeletal muscle: 1078).

Additionally, in any metabolic network, many reaction fluxes have a significant degree of flux correlation. This can arise from stochiometric coupling between reactions (e.g., the product of the first reaction is the substrate of the second reaction) and from proteins that mediate multiple reactions (e.g. some transmembrane carriers can transport a wide range of substrates). To account for this and facilitate the interpretation of the results, for each analyzed organ metabolic network, we pruned reaction fluxes with more than 50% correlation from the interaction results. Briefly, Pearson correlation coefficients were computed between all reaction fluxes from each organ. Next, all SNP-reaction fluxes pairs were ranked based on the maximum P-value between the interaction effect size test and the dosage-specific test. Then, starting from the reaction flux in the most significant reaction-SNP pair, reactions with more than 0.5 flux correlation (r) to this reaction were identified and all pairs involving these reactions were removed. The process was subsequently repeated for all ranked reaction-SNP pairs until no pairs involving reactions from the same organ-metabolic network with more than 0.5 flux correlation remained. In total 1,670 reactions remained (adipose tissue: 280, brain: 417, heart: 360, liver: 263, skeletal muscle: 350)

The interaction effect size test and the dosage-specific test P-values were adjusted for multiple testing against all remaining SNP-reaction flux pairs using the Benjamini and Hochberg (i.e., FDR) method.

Additionally, the univariate effect of the uncorrelated reactions on disease risk was also evaluated using a Cox model stratifying by sex and genotyping array and using the first 10 genetic principal components as covariates. The resulting p-values were FDR-adjusted for all uncorrelated reactions within each disease definition.

### Linkage disequilibrium estimation and variant annotation

LD between risk SNPs or SNPs used in flux prediction was measured using the genotype data from the UKB subset of inferred European ancestry using Plink 1.9^103^. LD blocks used to delimitate independent risk loci were defined using hierarchical clustering with the single linkage method (“friends of friends” clustering) using 1-R^2^ between risk variants as a distance metric. The resulting hierarchical tree was cut at a height of 0.4, identifying LD blocks such that each risk SNP had an R² > 0.6 with at least one other risk SNP within the block and an R² < 0.6 with all risk SNPs outside the block.

Ensembl Variant Effect Predictor was used to annotate the effect of risk variants involved in interactions^104^. Similarly, known eQTL and sQTL variants and their target genes were obtained from GTEx Portal (https://gtexportal.org/home/)^100^ and the INTERVAL RNA-SEQ Portal (https://www.intervalrna.org.uk/)^105^ and were used to further annotate the risk variants involved in interactions.

Gene features shown in plots were extracted from the TxDb.Hsapiens.UCSC.hg19.knownGene package with the makeGenesDataFromTxDb function from the karyoploteR package^106^.

## Data Availability

The data from UK Biobank is under restricted access as it contains potentially identifying and sensitive patient information. It can be accessed by making a reasoned request to UKB (https://www.ukbiobank.ac.uk/). All other data produced in the present study are available in the supplementary materials or upon reasonable request to the authors.

## Conflicts of Interest

A.S.B. reports institutional grants from AstraZeneca, Bayer, Biogen, BioMarin, Bioverativ, Novartis, Regeneron and Sanofi. J.D. serves on scientific advisory boards for AstraZeneca, Novartis, Our Future Health and UK Biobank, and has received multiple grants from academic, charitable and industry sources outside of the submitted work. M.I. is a trustee of the Public Health Genomics (PHG) Foundation, a member of the Scientific Advisory Board of Open Targets, and has research collaborations with AstraZeneca, Nightingale Health and Pfizer which are unrelated to this study.

## Acknowledgements

This research has been conducted using the UK Biobank Resource under Application 7439. This work was performed using resources provided by the Cambridge Service for Data-Driven Discovery (CSD3) operated by the University of Cambridge Research Computing Service (www.csd3.cam.ac.uk), provided by Dell EMC and Intel using Tier-2 funding from the Engineering and Physical Sciences Research Council (capital grant EP/P020259/1), and DiRAC funding from the Science and Technology Facilities Council (www.dirac.ac.uk). This work was supported by core funding from the British Heart Foundation (RG/18/13/33946: RG/F/23/110103), NIHR Cambridge Biomedical Research Centre (NIHR203312) [*], BHF Chair Award (CH/12/2/29428), Cambridge BHF Centre of Research Excellence (RE/18/1/34212), and by Health Data Research UK, which is funded by the UK Medical Research Council, Engineering and Physical Sciences Research Council, Economic and Social Research Council, Department of Health and Social Care (England), Chief Scientist Office of the Scottish Government Health and Social Care Directorates, Health and Social Care Research and Development Division (Welsh Government), Public Health Agency (Northern Ireland), British Heart Foundation and the Wellcome Trust. X.J. was also supported by the Wellcome Trust [227566/Z/23/Z]. M.I. was also supported by the UK Economic and Social Research 878 Council (ES/T013192/1).

*The views expressed are those of the authors and not necessarily those of the NIHR or the Department of Health and Social Care.

## Supplementary Figures and Tables

**Table S1:** Cases and controls under the definition of coronary atherosclerosis or myocardial infarction in the inferred European ancestry subset of UKB. Under the definition of coronary atherosclerosis, individuals with any of the constitutive ICD codes of ischemic heart disease were excluded from the controls.

**Figure S1:**
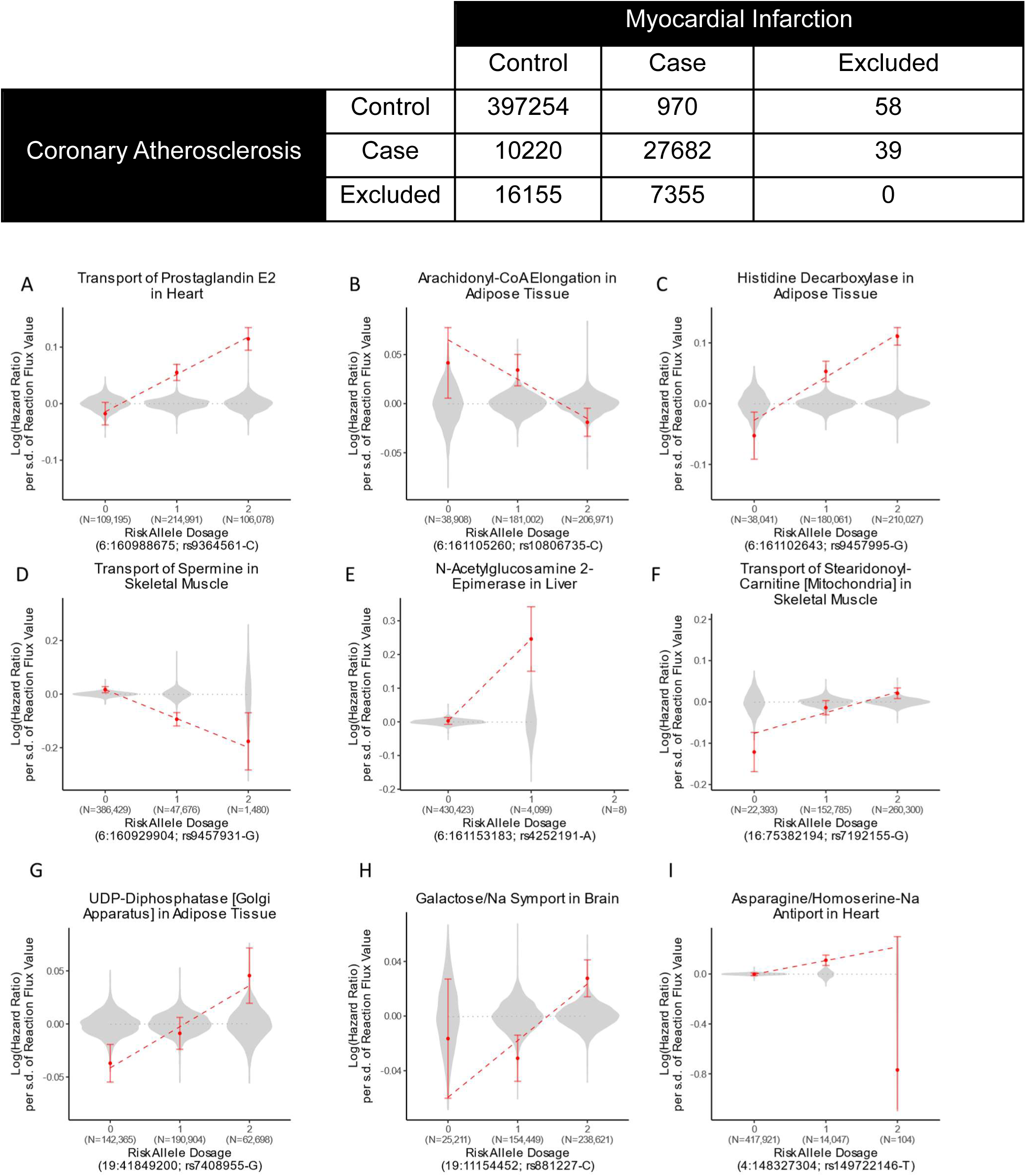
Examples of risk allele dosage-specific reaction effect sizes for a representative set of SNP-reaction flux pairs with significant interaction. Reaction flux effect sizes (i.e. log(Hazard ratio)) on CAD risk were estimated using Cox regression in European UKB participants carrying different risk allele dosages. Error bars denote the 95% confidence intervals for reaction effect sizes. The dashed line indicates the linear regression of flux effect size per dosage weighted by the standard error of effect size estimates (1/SE^2^). Violin plots indicate the distribution of all other risk reaction flux effect sizes for the same risk allele and the dotted line is the linear regression for these effect sizes. Genome coordinates correspond to the GRCh37 genome assembly.

**Figure S2:**
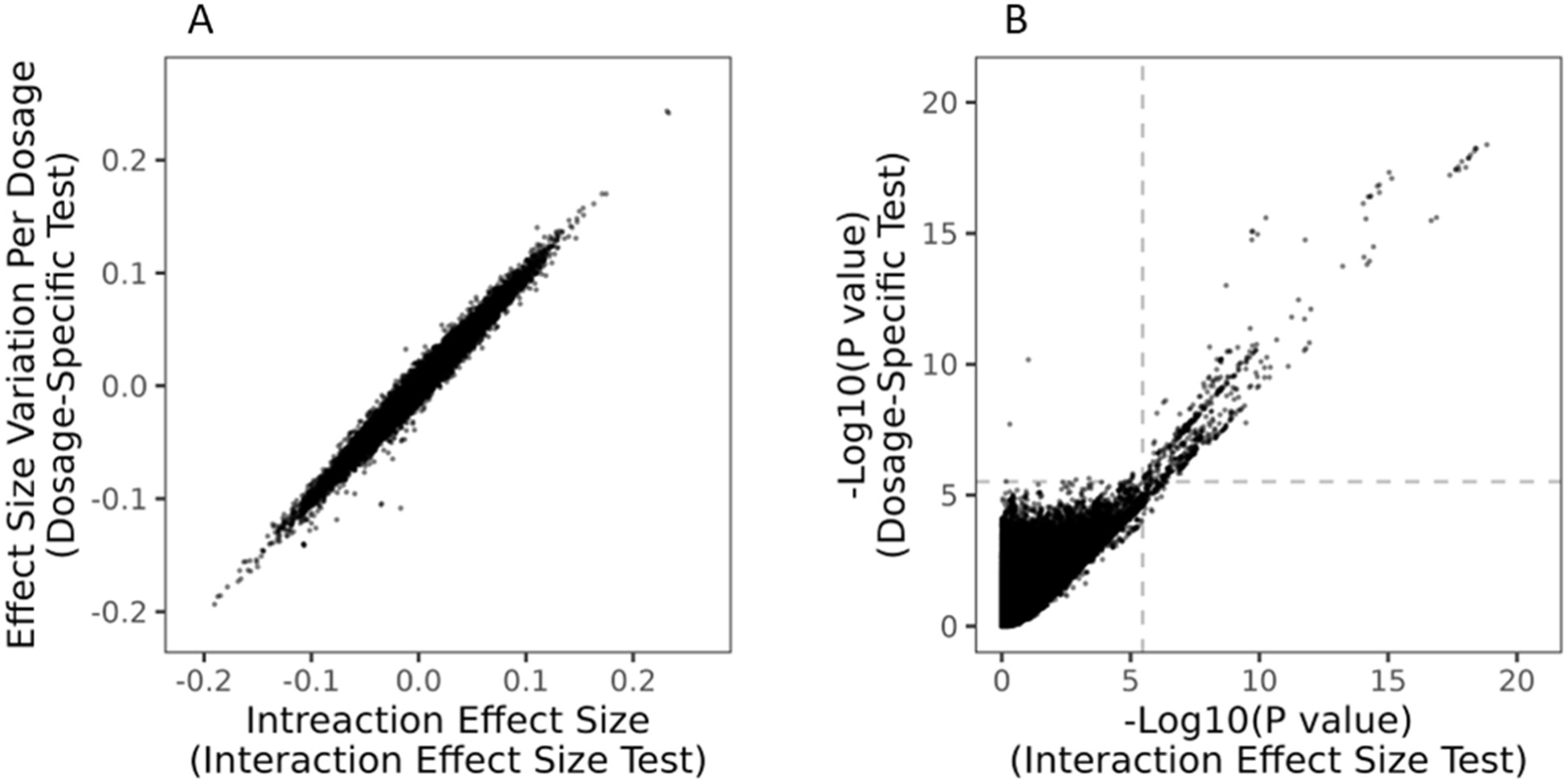
Comparison of interaction effect size and dosage-specific tests. A) Comparison of interaction effect sizes estimates. B) Comparison of interaction effect significance (-log10(P-values). The dashed grey line indicates the nominal P-value significance threshold estimated after Benjamini-Hochberg multiple testing correction (i.e., FDR<0.05).

**Figure S3:**
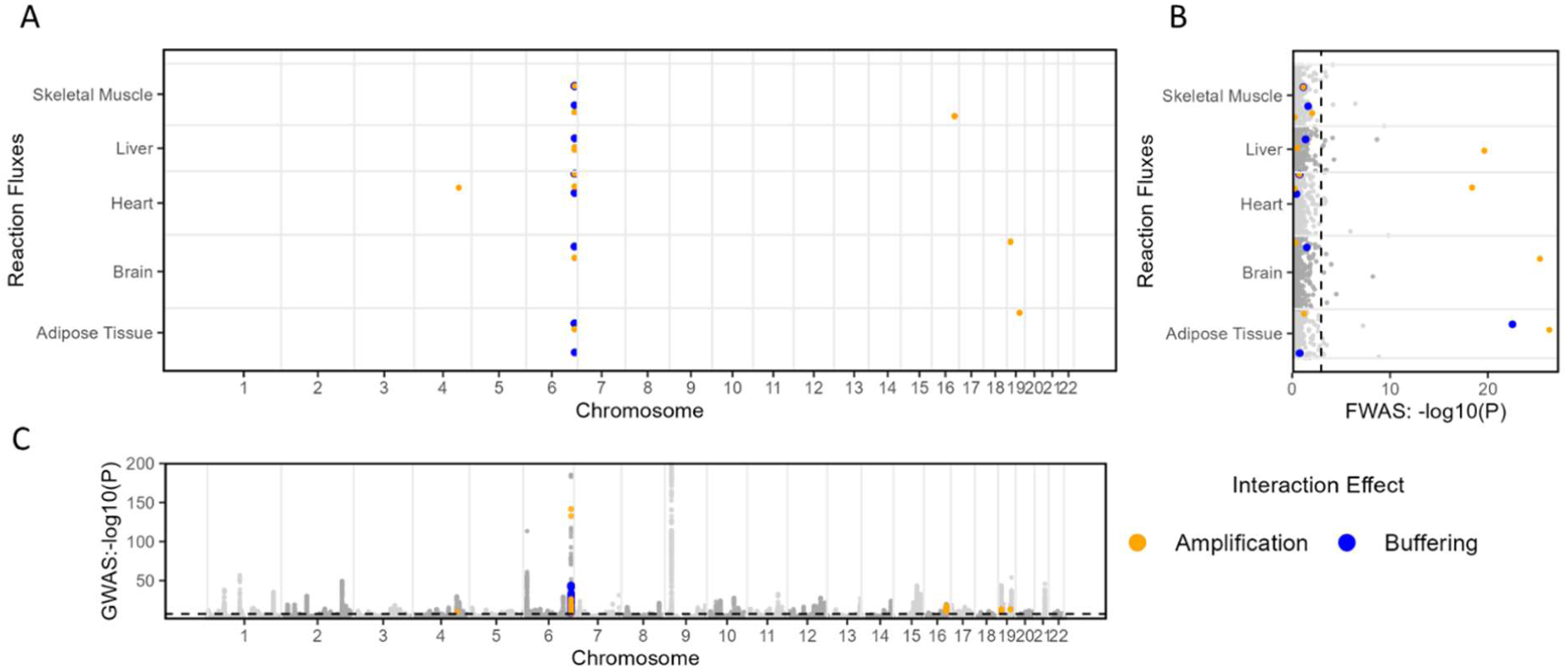
Variants and reaction fluxes with significant interaction. A) Organ metabolic network and chromosome of the reaction fluxes and variants, respectively, with significant interaction on CAD risk. B) and C) Manhattan plots of reaction fluxes and variants effects on CAD risk when analysed in univariate analysis. Variant effect sizes P-values were obtained from a published GWAS meta-analysis^28^. P-values for reaction effect size were computed for uncorrelated reaction fluxes with a Cox proportional hazard model in the inferred European ancestry subset of UKB. The dashed grey line indicates the P-value threshold to consider a variant or reaction effect size significant.

**Figure S4:**
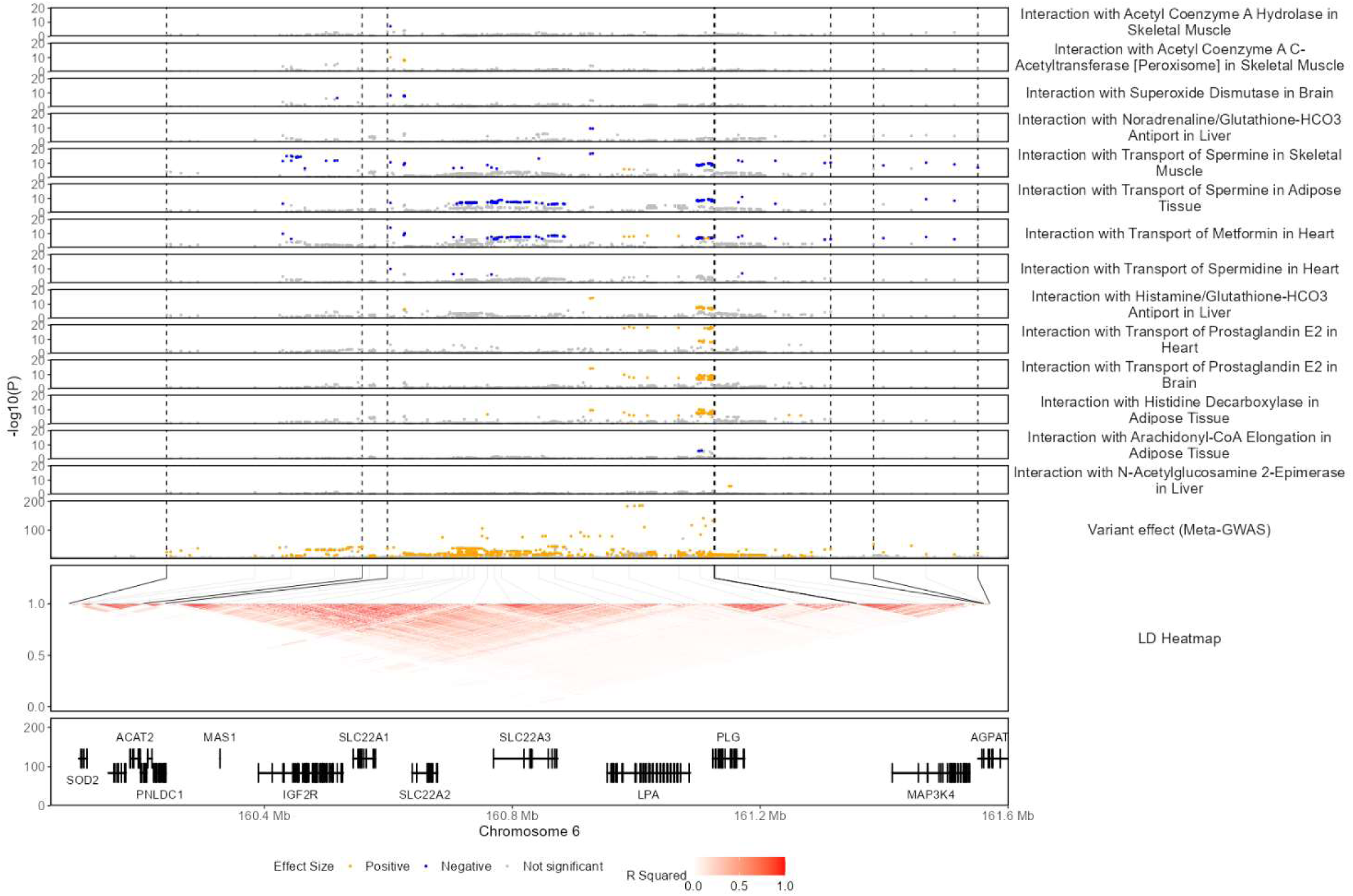
Significant SNP-Flux Interactions in the LPA/PLG risk loci. The regional association plots show the −log10(P value) for interaction and variant effect sizes on CAD risk. P-values for interaction and variant effect sizes were derived from the interaction effect size test and meta-GWAS summary statistics, respectively. The LD heatmap indicates the pairwise LD for SNPs with genome-wide significant effect size on CAD in UKB participants of European genetic ancestries. Dashed black lines indicate the limits of LD blocks (R^2^>0.6) used to define independent risk loci. To facilitate visualization, only LD blocks with variants involved in significant interactions are highlighted. Only protein-coding genes are shown in the gene plot. Genome coordinates correspond to the GRCh37 genome assembly.

**Figure S5:**
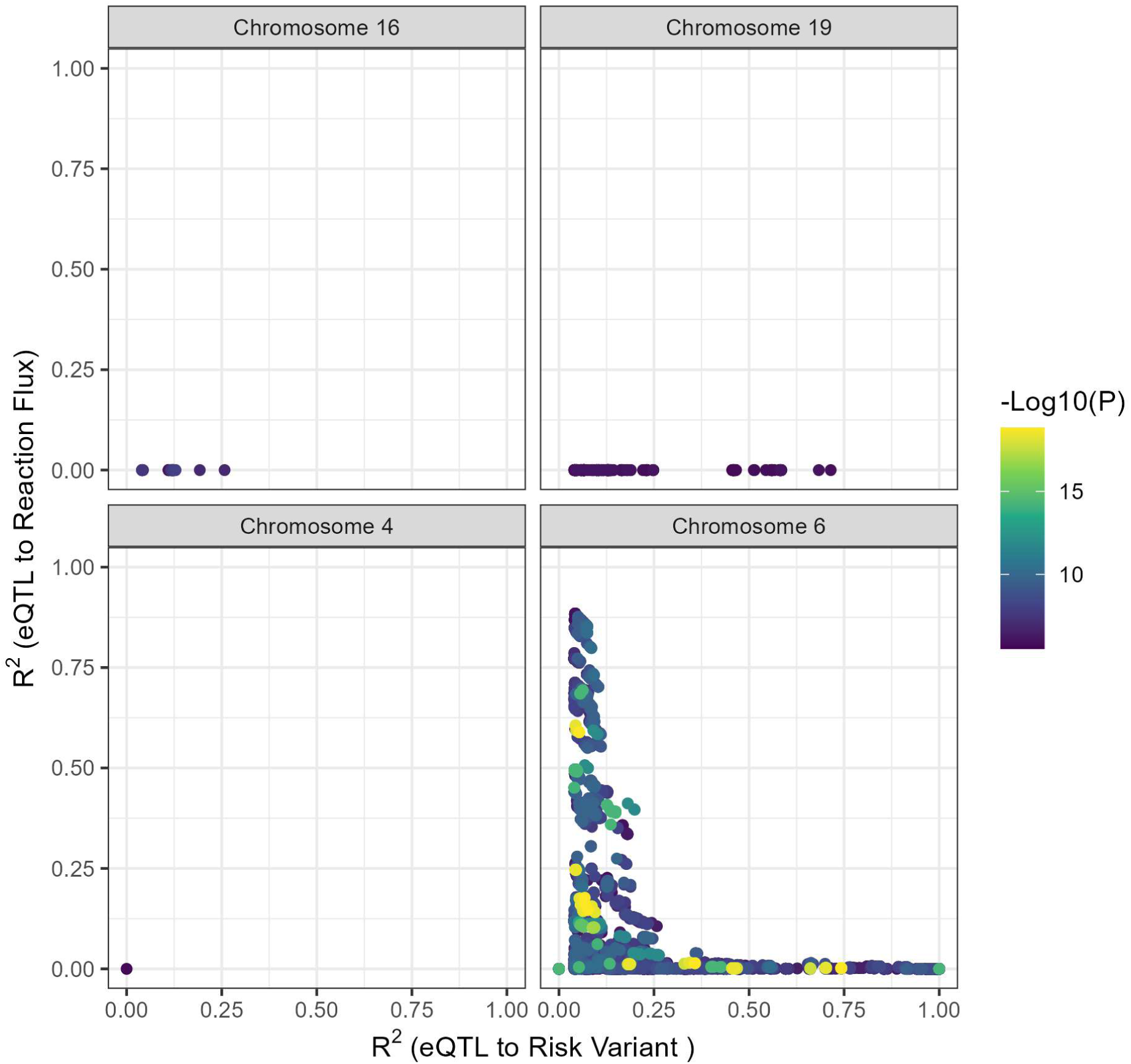
Linkage disequilibrium between risk variants and eQTL variants used as input for flux simulation and their correlation to the reaction flux value for each pair of risk-variant and reaction flux with significant interaction in CAD risk. For each pair, the effect of the risk variant has been regressed out of the flux (**Methods**). Linkage disequilibrium under 0.04 are plotted as 0. Data is coloured based on the interaction effect size P-value for each risk-variant reaction flux pair.

**Figure S6:**
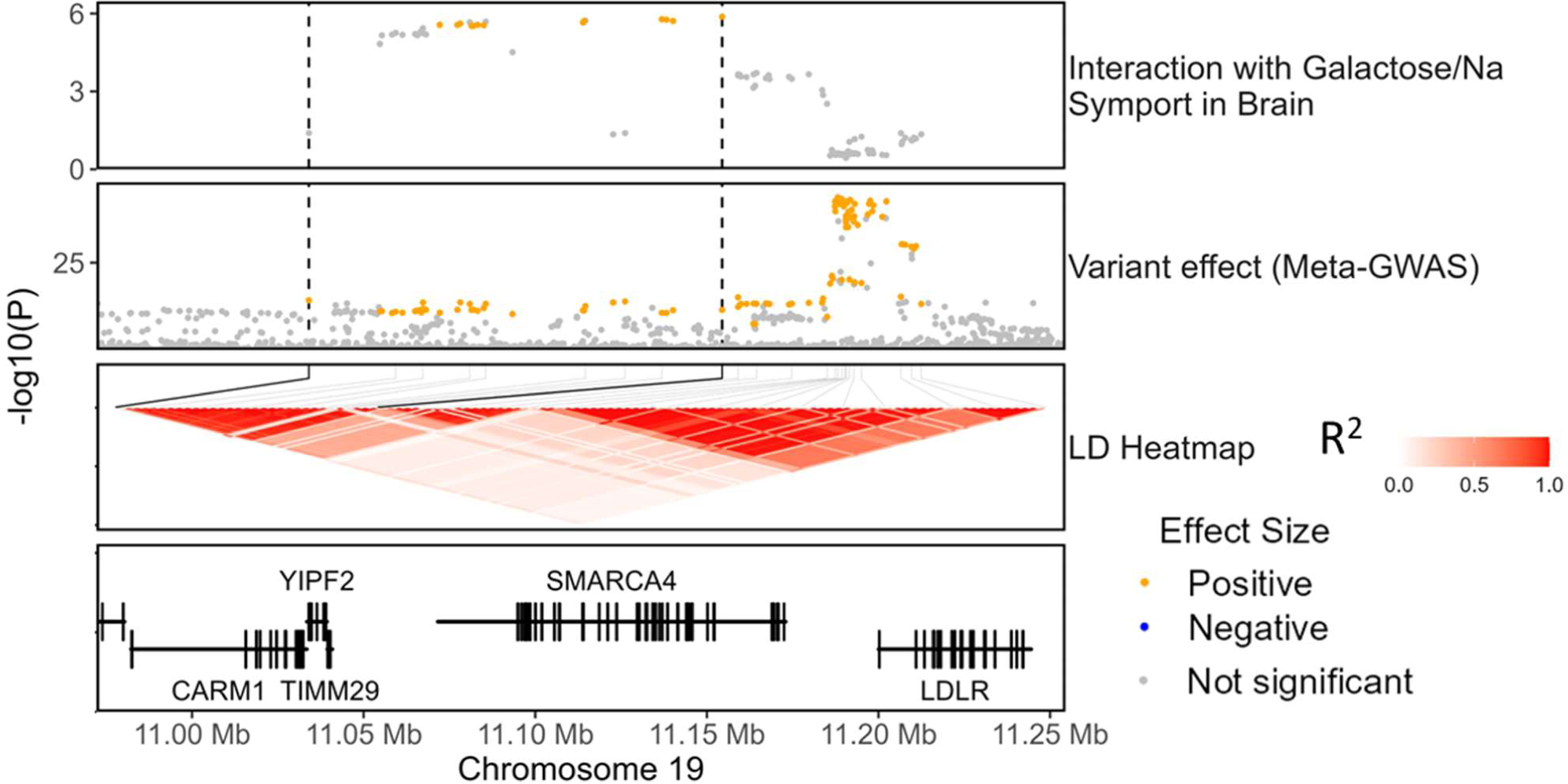
Interaction between galactose transport in brain and variants in the *SMARCA4* risk locus. The regional association plots show the −log10(P-value) for interaction and variant effect sizes on CAD risk. P-values for interaction and variant effect sizes were derived from the interaction effect size test and meta-GWAS summary statistics, respectively.

**Figure S7:**
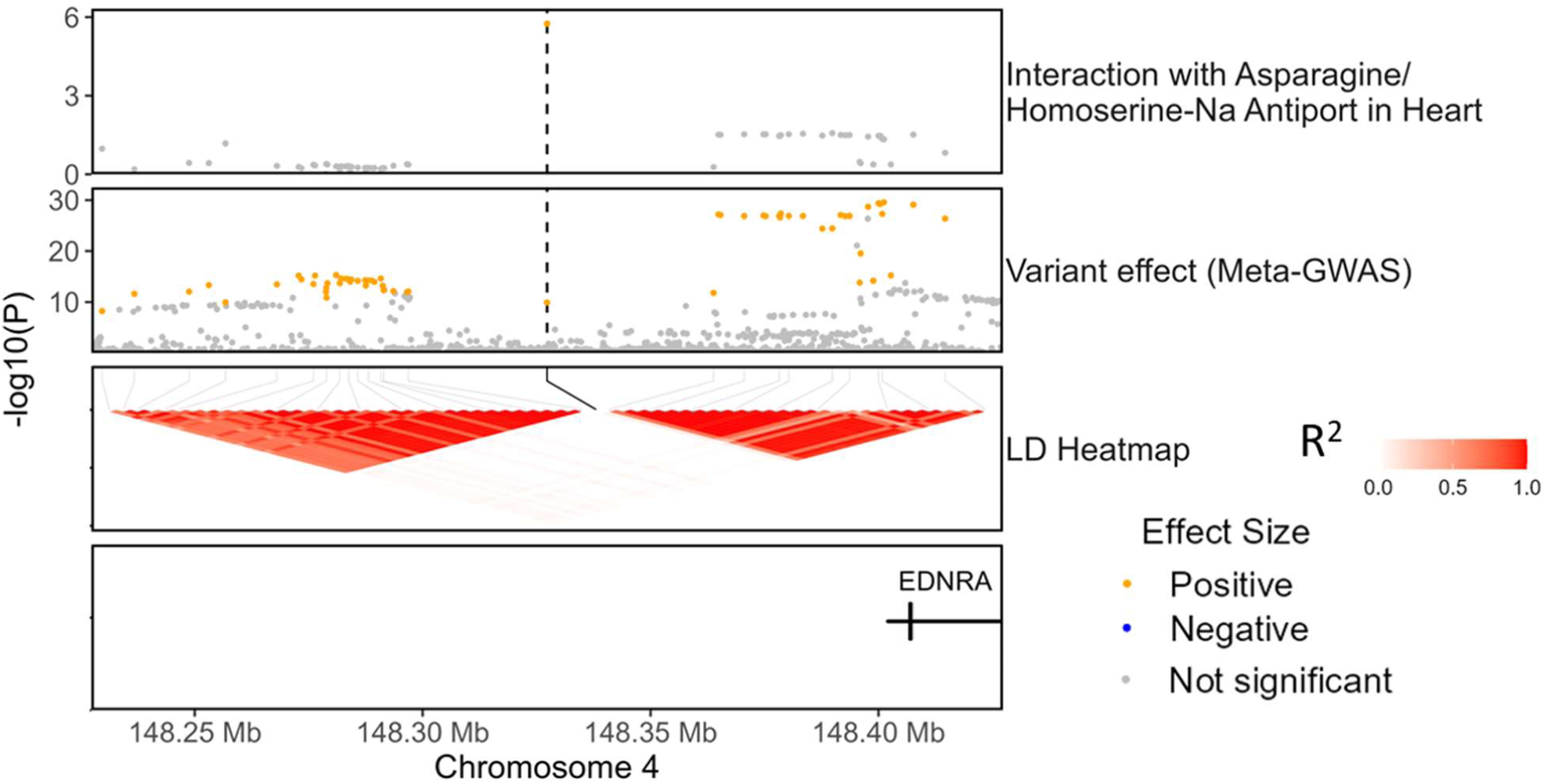
Amino acid transport amplifies the effect of a risk variant at the EDNRA locus. The regional association plots show the −log10(P-value) for interaction and variant effect sizes on CAD risk. P-values for interaction and variant effect sizes were derived from the interaction effect size test and meta-GWAS summary statistics, respectively.

**Figure S8:**
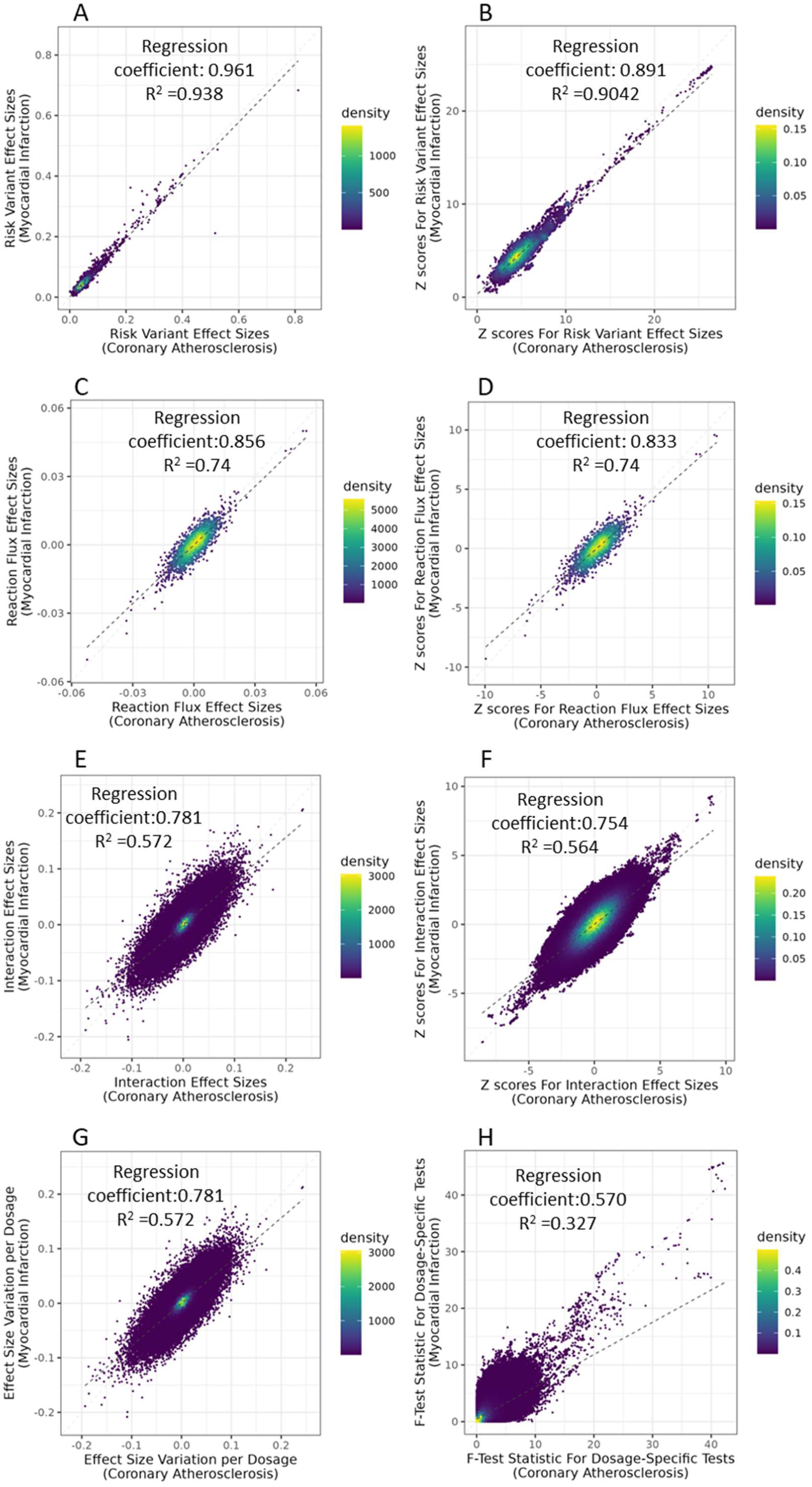
Effect sizes for the risk of coronary atherosclerosis and myocardial infarction. Effect sizes and Z scores for effect sizes for risk variants (A,B), reaction fluxes (C,B), interaction between reaction fluxes and risk variants (E,F), and reaction effect size variation and F-statistic for the dosage-specific test (G,H) using two alternative disease definitions: coronary atherosclerosis and myocardial infarction. The set of evaluated risk variants was obtained from a published GWAS meta-analysis^28^. The reaction flux effect size was evaluated for 1,670 uncorrelated reaction fluxes (**Methods**). Effect sizes were evaluated with a Cox proportional hazard model in the inferred European ancestry subset of UKB. The dosage-specific test evaluated the variation of reaction flux effect size per each risk allele dosage in the same UKB subset (**Methods**). The dashed black line indicates the linear regression for the plotted values.

**Figure S9:**
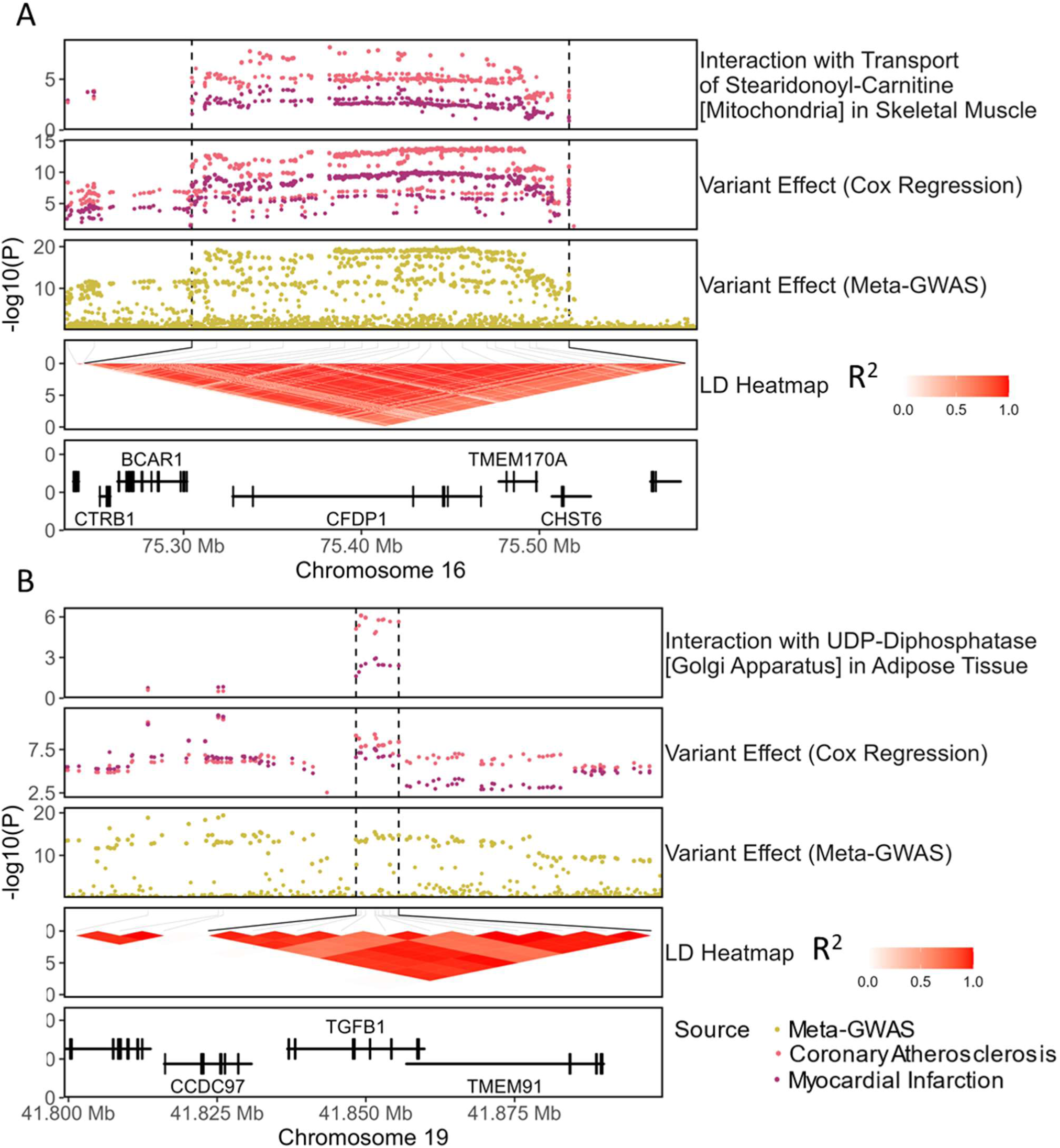
Example of interactions that are attenuated in myocardial infarction. The regional association plots show the −log10(P-value) for interaction and variant effect sizes on disease risk. P-values for interaction effect sizes and variant effect (Cox Regression) were derived from the interaction and variant effect size test, respectively, in the European subset of UKB for coronary atherosclerosis or myocardial infarction events. As a reference, the variant effects for the meta-GWAS on CAD risk are also plotted.

**Figure S10:**
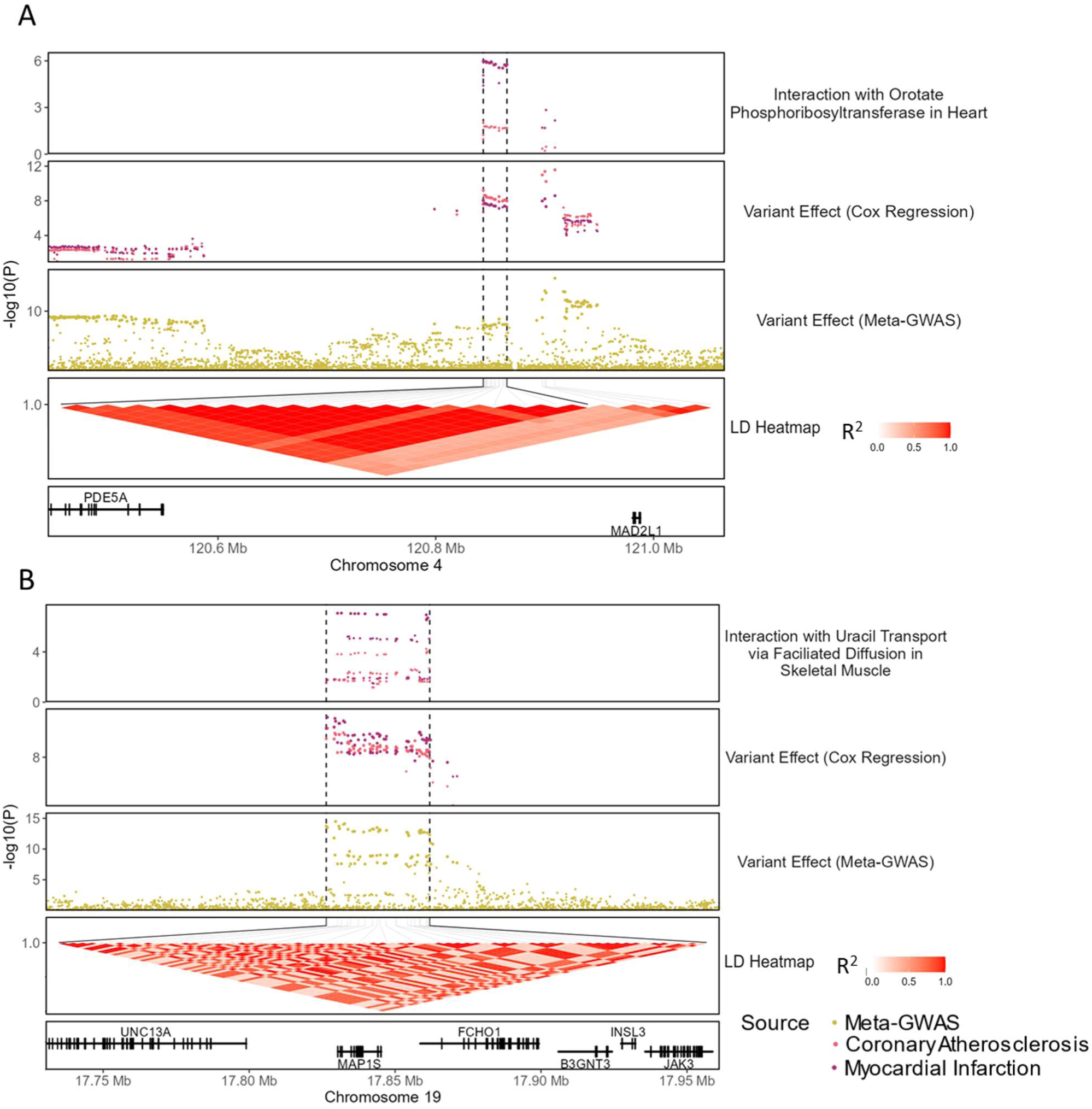
Example of interactions that are specific to myocardial infarction. The regional association plots show the −log10(P-value) for interaction and variant effect sizes on disease risk. P-values for interaction effect sizes and Variant Effect(Cox Regression) were derived from the interaction and variant effect size test, respectively, in the European subset of UKB for coronary atherosclerosis or myocardial infarction events. As a reference, the variant effects for the meta-GWAS on CAD risk are also plotted.

**Supplementary Data S1:** Table providing the reaction and variant annotation, and summary stats for pairs of risk alleles and reaction fluxes with significant interaction in either CAD or MI risk. Human1 and Recon3D Reaction IDs indicate the identifiers of the reactions in the HUMAN1 and Recon3D genome-scale models of human metabolism. Reaction organ indicates the organ-specific metabolic network where the flux is located. Genes mapped to reaction indicate the enzymes or transmembrane carriers mediating the reactions as defined in the gene-reaction rules of HUMAN1. eGene(s) and sGene(s) indicate the genes whose expression and splicing, respectively, are associated with the risk alleles. Variant risk locus indicates the LD block(R^2^>0.6) of risk variants where interacting risk alleles are mapped. The interaction between risk alleles and reaction fluxes is measured with two complementary approaches: an interaction effect size test and a dosage-specific test (**Methods**). Genome coordinates correspond to the GRCh37 genome assembly.

## Notes

### Author Declarations

This research has been conducted using the UK Biobank Resource under Application 7439. UK Biobank has approval from the North West Multi-centre Research Ethics Committee (Research Ethics Committee approval number: 21/NW/0157) as a Research Tissue Bank (RTB). This approval means that researchers do not require separate ethical clearance and can operate under the RTB approval.

